# Neurocognitive Impairment in Ugandan Children with Sickle Cell Anemia Compared to Sibling Controls: A cross-sectional study

**DOI:** 10.1101/2023.11.09.23298329

**Authors:** Paul Bangirana, Amelia K. Boehme, Annet Birabwa, Robert O. Opoka, Deogratias Munube, Ezekiel Mupere, Phillip Kasirye, Grace Muwanguzi, Maxencia Musiimenta, George Ru, Nancy S. Green, Richard Idro

## Abstract

**Introduction:** Neurocognitive function in Ugandan children aged 1-12 years with sickle cell anemia (SCA) were compared to their non-SCA siblings to identify risk factors for disease-associated impairment.

**Methods:** This cross-sectional neurocognitive function study of children with SCA (N=242) and non-SCA siblings (N=127) used age- and linguistically-appropriate standardized tests of cognition, executive function and attention for children ages 1-4 and 5-12 years. Test scores were converted to locally derived age-normalized z-scores. The SCA group underwent standardized stroke examination for prior stroke and transcranial doppler ultrasound (TCD) to determine stroke risk by arterial flow velocity.

**Results:** The SCA group was younger than siblings (mean ages 5.46±3.0 versus 7.11±3.51 years, respectively; p <.001), with lower hemoglobin concentration (7.32±1.02 vs. 12.06±1.42, p <.001). Overall cognitive SCA z-scores were lower: -0.73 ±0.98 vs. siblings -0.25 ±1.12 (p<.001), with comparable findings for executive function of -1.09±0.94 versus -0.84±1.26 (p=0.045), respectively. Attention z-scores for ages 5-12 for the SCA group and controls were similar: -0.37±1.4 vs. -0.11±0.17 (p=.09). Overall differences by SCA status were largely driven by the older age group, as z-scores in the younger sub-sample did not differ from controls. Analyses revealed the strongest predictors of poor neurocognitive outcomes among the SCA sample to be the disease, age and prior stroke (each p<.001). Impact from anemia and SCA were indistinguishable.

**Discussion:** Neurocognitive testing in children with SCA compared to non-SCA siblings revealed poorer SCA-associated functioning in children older than age 4. Results indicate need for trials assessing impact from disease modification for children with SCA.

## Introduction

Sickle cell anemia (SCA) is a serious inherited blood condition affecting 0.5-2% of births in Uganda and other high-prevalence countries in sub-Saharan Africa.^1-5^ High disease burden, compounded by health and health systems challenges in low-income countries, expose many affected children to early disease complications, including cerebrovascular injury.^4,6-9^ SCA-associated cerebrovascular injury commonly results in overt and/or clinically “silent” infarcts, often in children under age 10 years.^10-14^ Infarcts can lead to impaired neurocognitive function.^15-18^ In high-income countries where successful stroke prevention strategies are routinely practiced, continued occurrence of silent infarcts remains a neurocognitive risk.^10-12,18-21^ Worldwide, children with SCA have heightened risk of intellectual deficits with or without imaging abnormalities.^15,22^

Severe anemia is a risk factor for SCA-associated cerebral infarcts and impaired neurocognition due to abnormal blood flow and reduced cerebral oxygen delivery.^19,23-28^ Risk of cognitive impairment from SCA in sub-Saharan Africa may be compounded by low parental education, a proxy for poverty, malnutrition and endemic infections.^29-34^ Moreover, stroke reduction strategies are not generally available in the region.^14,35,36^ Cerebrovascular injury among the many African children with SCA raise questions about prevalence and types of neurocognitive risk in this population.^36^ To date, few pediatric studies of SCA in sub-Saharan Africa have assessed the associated neurocognitive effects compared to unaffected children.^27,29,30,37^ Only one of these studies compared results to sibling controls, a strategy which can better control for environmental and socio-economic affects.^27^

We assessed the frequency of neurological and neurocognitive impairment in a cross-sectional study of Ugandan children with SCA ages 1-12 years, “Burden and Risk of Neurological and Cognitive Impairment in Pediatric Sickle Cell Anemia in Uganda (BRAIN SAFE).”^14^ Overall frequency of neurocognitive dysfunction was 11.2%, with older (ages 5-12 years) at a 3-fold higher risk of impairment compared to younger participants (ages 1-4 years). In this secondary analysis, we report detailed findings of neurocognitive evaluation of participants compared to their non-SCA siblings to identify contributions from demographic and clinical factors beyond age. We hypothesized that, compared to non-SCA siblings, children with SCA had lower neurocognitive function and that age, malnutrition and adverse neurological outcomes of prior stroke and elevated TCD velocity were risk factors. In contrast to other sub-Saharan Africa studies of children with SCA, we assessed detailed neurocognitive performance for cognition, executive function and attention in a large sample of Ugandan children compared to non-SCA siblings, as well as the effects of key demographic and neurological risk factors.

## Materials and Methods

### Study design and setting

A random cross-sectional sample of 265 children with SCA ages 1-12 years attending the Mulago Hospital Sickle Cell Clinic in Kampala, Uganda and a sample of their non-SCA siblings were enrolled in BRAIN SAFE 1 (2016-2018).^14^ Sample size was determined from previously reported frequency and impact of cerebral infarction on neurological and neurocognitive function.^15-18^ Routine SCA pediatric care did not include disease-modifying therapy at that time. The study was approved by Makerere University School of Medicine Research and Ethics Committee, Uganda National Council for Science and Technology and Columbia University Institutional Review Board.

### Participants

As previously reported, inclusion criteria were: a) SCA confirmed by hemoglobin electrophoresis (HbSS or HbS-B^0^ thalassemia); b) having attended the Mulago SCA clinic.^14^ To focus on SCA-related neurological complications, we excluded those with a history of neurological abnormalities before 4 months of age.^38^ Caregiver written informed consent was obtained, with assent from participants aged eight years or older. Non-SCA participants (N=127) were also enrolled, with inclusion criteria of: a) aged 1-12 years; and b) hemoglobin electrophoresis demonstrating lack of SCA (i.e. HbAA or HbAS). Among these controls, 119 (93.7%) were siblings; the rest were other close relatives or neighbors. Hence, we refer to them as “siblings.”

### Physical and Neurological assessments and Caregiver Education

Anthropometric assessments of malnutrition for the SCA and sibling participants for detecting malnutrition defined as low weight-for-height (“wasting”) used World Health Organization (WHO) standards, as previously reported. ^14, 39^ Assessments of SCA participants at enrolment were: medical history and physical examination, examination for prior stroke using NIH Pediatric Stroke Scale (PedNIHSS) and stroke risk stratification by intracranial arterial flow velocity identified by transcranial doppler (TCD) as elevated to ≥170cm/sec (“conditional” or “abnormal”).^14^ Caregiver educational attainment was scored as previously performed, reported as none, primary school, secondary school, more than secondary education or unknown.^40^

### Neurocognitive assessment

Overall neurocognitive function, including behavioral measures, attention and executive function were assessed using age-appropriate tests by experienced testers in both SCA and non-SCA siblings. All assessment tools had previously been translated into a predominant local language and used to establish age-specific community norms for healthy children in Kampala.^14^

For children aged 1-4 years, the Mullen Scales of Early Learning (Mullen) ^41^ and the Behavioral Rating Inventory for Executive Function-Preschool version (BRIEF-P) assessed cognitive functioning and executive function, respectively.^42^ The Mullen sub-tests assess gross and fine motor, visual reception, receptive language and expression language. Summation of fine motor, visual reception, receptive language and expressive language scores constitute the Early Learning Composite to measure overall neurocognitive ability, the primary outcome for the Mullen. The BRIEF-P is a caregiver assessment of the child’s executive functioning using 63 items for which the caregiver endorses child behaviors exhibited over the prior six months. Summation of these items gives the Global Executive Composite to measure executive function, the primary outcome of the BRIEF-P. Sub-tests were for self-control, flexibility and metacognition.

Children aged 5-12 years were tested using the Kaufman Assessment Battery for Children, second edition (KABC-II),^43^ BRIEF school-age version^44^ and Test of Variables of Attention (TOVA)^45^ to assess overcall neurocognitive functioning, executive function and attention, respectively. KABC-II subscales assessed working memory, visual spatial ability, learning ability and reasoning. Summation of these four scales generates the composite value, the Global Mental Processing Index. The BRIEF for school-age participants uses caregiver responses on 86 items. Here, the composite score, computed from the subtests of behavioral regulation and metacognition, generated the General Executive Composite. The TOVA, a computerized test for which children are instructed to press a switch whenever a specific target appeared on the screen, assesses attention and inhibitory control. The composite score, D’ Prime, is calculated from subtest scores for omission errors, commission errors, response time and attention-deficit/hyperactivity disorder (ADHD).

### Statistical analyses

SCA and non-SCA participants were grouped by the two age ranges according to the tests used. Raw scores for all neurocognitive assessments were converted to age-normalized z-scores using established standards for unaffected healthy children, as previously described.^14^ Within each age range, z-scores were analyzed and compared, by group, using means and standard deviations. Negative values correspond to scores below the age-normalized z-scores. In contrast, negative z-scores for BRIEF and BRIEF-P (pre-school) indicate better function. Hence the positive signs for those two BRIEF tests were flipped to negative for consistent directionality for reporting results.^46^ Data were analyzed by means and standard deviation. For categorical data, ANOVA and Pearson chi-square was used for analyses. Continuous data were analyzed by Pearson correlation. Factors associated with impaired neurocognition were assessed using linear regression. Missing data were removed.

## Results

### SCA and Non-SCA Siblings Demographic and Clinical Characteristics

Neurocognitive assessment was performed in 242 of 265 (91.3%) SCA participants and all 127 non-SCA siblings. Parents of the 23 SCA participants without neurocognitive assessments were unable to schedule testing.^14^ Mean age of SCA participants tested was 5.46±2.98 years versus 7.11±3.51 in the non-SCA siblings (p<.001), (Table 1). Mean hemoglobin concentration was also highly different by group: 7.32±1.02 versus 12.06±1.42 in the SCA versus control group, respectfully (p<.001). Close to half (48.4%) of the SCA sample was female compared to 43.9% of the non-SCA siblings (p=.33). Malnutrition, defined as weight-for-age at -2 z-scores or below (wasting) using World Health Organization (WHO) global norms by age and sex, was found in 37 (15.3%) of the children with SCA and 11 (8.7%) of the controls (p=.20). Caregiver education differed between the two study groups (p=.018), with a higher proportion of caregivers in the control group having little or no education.

**Table 1.**
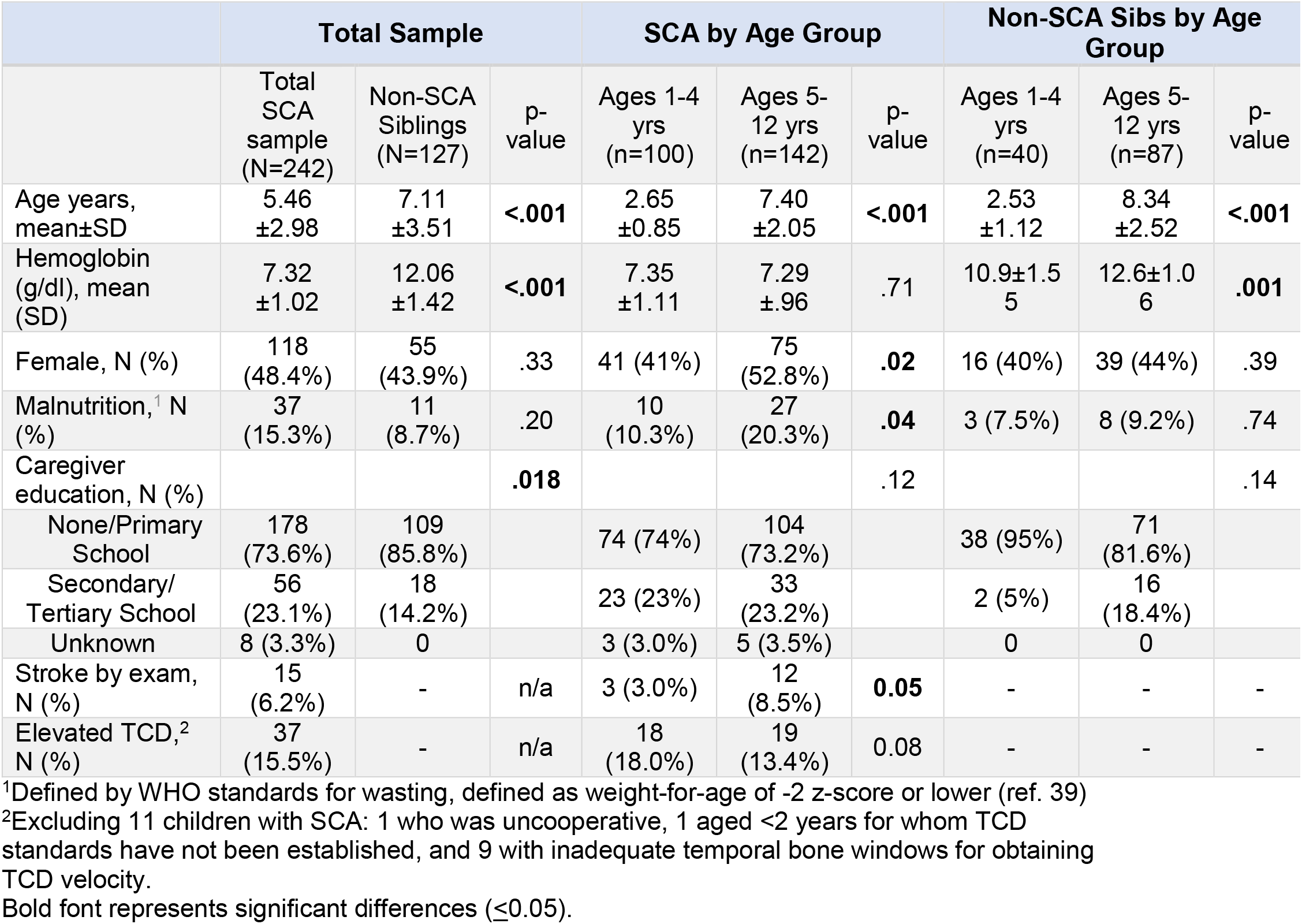
Demographic and neurologic characteristics stratified by SCA status and age group, 1-4 or 5-12 years of age. Data are expressed as N (%) unless otherwise stated.

|  | Total Sample |  |  | SCA by Age Group |  |  | Non-SCA Sibs by Age Group |  |  |
| --- | --- | --- | --- | --- | --- | --- | --- | --- | --- |
|  | Total SCA sample (N=242) | Non-SCA Siblings (N=127) | p-value | Ages 1-4 yrs (n=100) | Ages 5-12 yrs (n=142) | p-value | Ages 1-4 yrs (n=40) | Ages 5-12 yrs (n=87) | p-value |
| Age years, mean±SD | 5.46 ±2.98 | 7.11 ±3.51 | **<.001** | 2.65 ±0.85 | 7.40 ±2.05 | **<.001** | 2.53 ±1.12 | 8.34 ±2.52 | **<.001** |
| Hemoglobin (g/dl), mean (SD) | 7.32 ±1.02 | 12.06 ±1.42 | **<.001** | 7.35 ±1.11 | 7.29 ±.96 | .71 | 10.9±1.5<br>5 | 12.6±1.0<br>6 | **.001** |
| Female, N (%) | 118 (48.4%) | 55 (43.9%) | .33 | 41 (41%) | 75 (52.8%) | **.02** | 16 (40%) | 39 (44%) | .39 |
| Malnutrition,<sup>1</sup> N (%) | 37 (15.3%) | 11 (8.7%) | .20 | 10 (10.3%) | 27 (20.3%) | **.04** | 3 (7.5%) | 8 (9.2%) | .74 |
| Caregiver education, N (%) |  |  | **.018** |  |  | .12 |  |  | .14 |
| None/Primary School | 178 (73.6%) | 109 (85.8%) |  | 74 (74%) | 104 (73.2%) |  | 38 (95%) | 71 (81.6%) |  |
| Secondary/ Tertiary School | 56 (23.1%) | 18 (14.2%) |  | 23 (23%) | 33 (23.2%) |  | 2 (5%) | 16 (18.4%) |  |
| Unknown | 8 (3.3%) | 0 |  | 3 (3.0%) | 5 (3.5%) |  | 0 | 0 |  |
| Stroke by exam, N (%) | 15 (6.2%) | - | n/a | 3 (3.0%) | 12 (8.5%) | **0.05** | - | - | - |
| Elevated TCD,<sup>2</sup> N (%) | 37 (15.5%) | - | n/a | 18 (18.0%) | 19 (13.4%) | 0.08 | - | - | - |

<sup>1</sup>Defined by WHO standards for wasting, defined as weight-for-age of -2 z-score or lower (ref. 39)
<sup>2</sup>Excluding 11 children with SCA: 1 who was uncooperative, 1 aged <2 years for whom TCD standards have not been established, and 9 with inadequate temporal bone windows for obtaining TCD velocity.
Bold font represents significant differences (≤0.05).

Compared within each sample by age group, ages 1-4 versus 5-12 years, beyond significant differences by age, only the older control group had significantly higher hemoglobin concentration (Table 1). For the SCA sample, the older group had higher proportion of females and more malnutrition compared to the young group. Among neurological outcomes in the SCA sample, a higher proportion of the older group had a prior stroke and a marginally lower proportion with elevated TCD velocity.

### Overall Neurocognition in children with SCA vs. non-SCA siblings

Mean scores for the groups with SCA and non-SCA siblings were normally distributed for all three neurocognitive domains tested. The overall sample with SCA performed significantly worse for cognition in standardized age-appropriate tests than the non-SCA controls, -0.73±.98 vs. -0.25±1.12 (p<.001) (Table 2, Figure). Similarly, the SCA sample scored lower for executive function than the unaffected siblings: -1.09±0.94 vs. -0.84±1.26 (p=.045), respectively).

**Figure.**
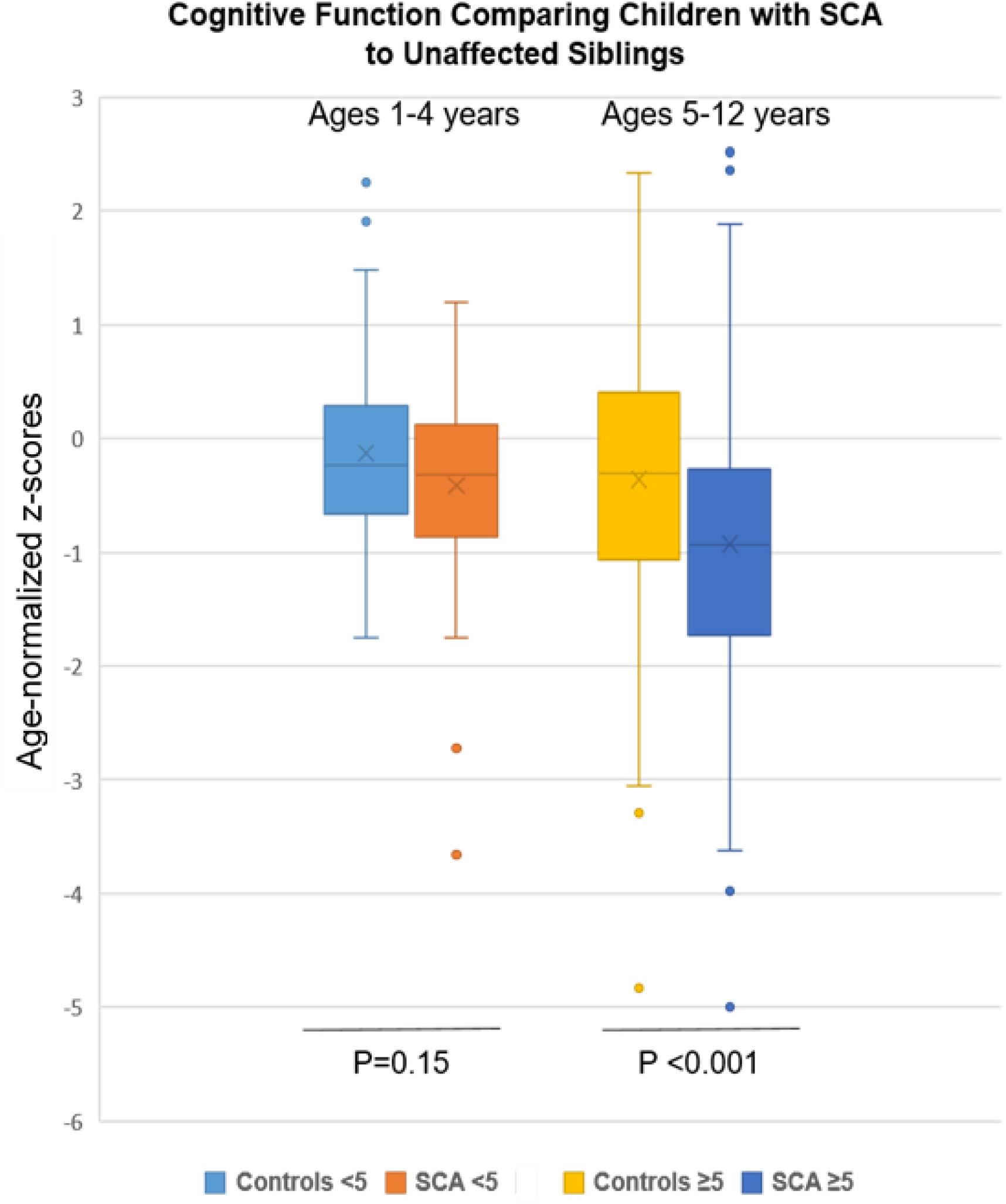
Cognitive findings by SCA status and age group (1-4 and 5-12 years) compared to unaffected siblings. Age-normalized z-scores for mean cognitive testing were lower in the SCA group, but only in the older age group (**p<.001**).

**Table 2.** Composite Neurocognitive test results, by z-scores, for Children Aged 1-4 and 5-12 Years, Assessed by Tests for Cognition, Executive Function and Attention (the last test was used only with the older group). Standardized age-normalized z-scores were scored based on local unaffected controls.^1^ (Ref. 14, 40)

| **Overall Differences by Sickle Cell Status, Aged 1-12 years** |  |  |  |
| --- | --- | --- | --- |
|  | SCA Participants (N=242) | Non-SCA Siblings (N=127) |  |
|  | Mean±SD (Range) | Mean±SD (Range) | p-value |
| Overall Cognition | -0.73±0.98 (-5.0-2.51) | -0.25±1.12 (-3.84-3.42) | **<.001** |
| Overall Executive Function | -1.09±0.94 (-2.56-3.25) | -0.84±1.26 (-2.87- 3.35) | **.045** |
| **Differences by Sickle Cell Status, Aged 1-4 Years** |  |  |  |
|  | SCA Participants (N=100) | Non-SCA Siblings (N=40) |  |
|  | Mean (SD) (Range) | Mean (SD) (Range) |  |
| Mullen Scales of Early Learning | -0.41±0.73 (-3.66, 1.20) | -0.21±0.84 (-1.65-3.42) | .15 |
| BRIEF-P: Global Executive function<sup>2</sup> | -1.47±0.89 (-1.01-3.25) | -1.61±1.22 (-2.87-3.35) | .44 |
| **Differences by Sickle Cell Status, Aged 5-12 years** |  |  |  |
|  | SCA Participants (N=142) | Non-SCA Siblings (N=87) |  |
|  | Mean (SD) (Range) | Mean (SD) (Range) |  |
| KABC-II mental processing index | -0.93±1.25 (-5.00-2.52) | -0.36 ±1.24 (-3.84-3.16) | **<.001** |
| BRIEF: Global Executive function<sup>2</sup> | -0.69±0.83 (-2.56-2.05) | -0.32±0.99 (-2.81-1.86) | **.009** |
| TOVA: D' Prime<sup>3</sup> | -0.37±1.4 (-3.31-1.80) | -0.11±0.17 (-3.10- 5.00) | .09 |

<sup>1</sup>The 2 to 5 component sub-tests for each neurocognitive test are listed in the Methods.
<sup>2</sup>BRIEF-P and BRIEF scores were flipped from positive to negative for the for consistent data directionality.
<sup>3</sup>This test is not valid in children below age 5.
Bold font represents statistically significant differences ( $p \leq 0.05$ ).

Among the younger age group tested, ages 1-4 years, no differences by SCA status were found in cognition or executive function. Cognitive function tested by the Mullen was -0.41±0.73 for SCA and -0.21±0.84 (p=.15) non-SCA samples. Similarly, executive function tested by the BRIEF-P in these younger participants comparing SCA vs. non-SCA was -1.47±0.89 vs. -1.61±1.22 (p=0.44), respectively. Hence, neurocognitive function was retained in the younger sub-sample of children with SCA through age 4 years.

In contrast, lower z-scores were found by SCA status for the older children for neurocognition tested by KABC-II: -0.93±1.25 vs. -0.36 ±1.24 (p<.001) and for executive function tested by the BRIEF: -0.69±0.83 vs. -0.32±0.99 (p=.009). However, testing for attention by TOVA, possible only among the older age group, demonstrated similar performance between the two groups: -0.37±1.4 versus -0.11±0.17 (0.09) for the SCA and control groups, respectively. Two of the three areas of assessment, cognition and executive function, were lower in the older sub-sample of children with SCA compared to controls.

To remove potential excess influence on cognition from prior stroke within the SCA sample, we re-analyzed mean z-scores after removing the 15 affected SCA participants (mean age 6.0±2.59 years). As expected, the mean z-score for the SCA sub-sample with prior stroke versus no stroke was much lower -2.18±1.53 than the overall scores (p<.001). However, removing the small subset affected by stroke from the SCA sample had no significant effects on the mean SCA z-scores for cognition or executive function compared to controls.

### Factors Associated with Impaired Cognition in SCA children vs. non-SCA siblings

Overall neurocognitive test results for each of the domains for SCA were compared to controls for each variable collected. We first asked whether hemoglobin concentration for the SCA group compared to controls had effects that were separable from SCA in neurocognitive outcomes. By linear regression using both variables in the model - SCA status and hemoglobin concentration - the difference was p=.004, with the main effect largely driven by SCA. No effects from sex, malnutrition or elevated TCD velocity were found in any of the three neurocognitive domains tested (Table 3).

**Table 3.** Neurocognitive outcomes compared between the groups with SCA and controls. By linear regression, adjusting for age, hemoglobin, caregiver education and prior stroke changed affect sizes but not outcomes of cognitition or executive function, thereby identifying SCA as the primary source of group differences.* In contrast, attention was largely influenced by SCA and age differences between the two samples.

| **Cognition** | **F Value** | **P-Value** |
| --- | --- | --- |
| Unadjusted | 16.1 | **<.001** |
| Model 1: Adjusted for age | 15.0 | **<.001** |
| Model 2: Adjusted for age, hemoglobin, caregiver education | 8.5 | **<.001** |
| Model 3: Adjusted for age, hemoglobin, caregiver education, prior stroke | 12.6 | **<.001** |
| **Executive Function** |  |  |
| Unadjusted | 10.8 | **.001** |
| Model 1: Adjusted for age | 5.4 | **.005** |
| Model 2: Adjusted for age, hemoglobin, caregiver education | 2.6 | **.034** |
| Model 3: Adjusted for age, hemoglobin, caregiver education, prior stroke | 2.3 | **.04** |
| **Attention** |  |  |
| Unadjusted | 2.9 | .09 |
| Model 1: Adjusted for age | 3.6 | **.03** |
| Model 2: Adjusted for age, hemoglobin, caregiver education | 2.0 | .10 |
| Model 3: Adjusted for age, hemoglobin, caregiver education, prior stroke | 2.6 | **.028** |

\*These variables had no effect on outcomes: sex, malnutrition, elevated TCD velocity. Caregiver education had a modestly significant impact on executive function.
<sup>1</sup> Hemoglobin concentration did not impact the variance beyond that from SCA.

We then examined overall test outcomes for factors contributing to the outcomes (Table 3). For overall cognition or executive function, age-normalized z-scores declined with age. Adjusting for other variables of hemoglobin concentration, caregiver education and prior stroke demonstrated their impact on effect sizes. Despite those changes, the outcomes were unchanged. These findings demonstrated that poorer function of the SCA group attributed to the impact of the disease.

By the TOVA test for attention in the older participants in both SCA and controls, age also negatively impacted z-scores. The influence of age for the SCA group was assessed as r^2^ = -.58 and -.72 for the controls (both p<.-001). Unexpectedly, neither prior stroke nor elevated TCD was associated with reduced attention.

Finally, we examined the SCA sample for impact from prior stroke or elevated TCD velocity on each of the three outcomes. Prior stroke strongly affected cognition (<.001), but had no significant effects on executive function or attention (Table 4). Elevated TCD velocity had a borderline impact on cognition but it, like prior stroke, had no impact on executive function or attention.

**Table 4.** Examining the role of prior stroke or elevated TCD velocity on neurocognitive outcomes within the SCA sample. By linear regression of age-normalized z-scores, adjusting for prior stroke demonstrated a strong relationship with cognition but not the other outcomes. In contrast, adjusting for elevated TCD velocity demonstrated that no outcome was affected.*

| **Prior Stroke** | F value | P-value |
| --- | --- | --- |
| Cognition | 25.1 | **<.001** |
| Executive Function | .06 | .43 |
| Attention | .14 | .71 |
| **Elevated TCD Velocity** |  |  |
| Cognition | 3.4 | .067 |
| Executive Function | .23 | .63 |
| Attention | .01 | .91 |

\*1 participant had both prior stroke and elevated TCD velocity.

## Discussion

Children with SCA in sub-Saharan Africa are at risk for disease-associated cerebrovascular injury as well as environmental challenges.^6,47^ In a large clinic-based sample of Ugandan children with SCA in Kampala compared to their non-SCA siblings, our cross-sectional neurocognitive assessment revealed these main findings: 1) Mean test z-scores for cognition and executive function were substantially lower in the SCA sample, even after accounting for age, hemoglobin concentration, caregiver education or prior stroke. No discernable effects were seen from sex, malnutrition or elevated TCD velocity. These findings confirm SCA as the main cause of impaired neurocognition in this study. Lower scores in cognition testing of approximately 0.5 z-scores in the children with SCA correspond to approximately 8 IQ points below that of the siblings; 2) Differences by hemoglobinopathy diagnosis were driven by the older sub-sample with SCA, ages 5-12 years.^48^ In contrast, children with SCA aged 1-4 years were not different in cognition or executive function from unaffected siblings in that age group; 3) Age and clinically evident prior stroke, but not elevated TCD arterial velocity, were the largest drivers of impaired cognition. Nonetheless, exclusion from the analyses of the modest number of SCA participants with prior stroke did not significantly affect the results of the remaining sample; 4) Among those aged 5-12 years, attention was not significantly different from the controls unless accounting for difference by age and/or prior stroke; 5) Non-SCA siblings in our study scored below healthy, Kampala-based age-normalized controls. What social, economic and/or educational opportunities affected neurocognitive performance in the siblings were not assessed.

Similar findings of impaired neurocognitive function and attention compared to non-SCA siblings were observed in a prior Tanzanian report of a smaller sample of older SCA children.^27^ Similar to prior reports of children with SCA in Africa, the U.S. and elsewhere, our study’s participants with SCA had lower executive function.^15,27,37^ This consistent finding was seen despite our use of a test by parental report which may under-report functional deficits, rather than direct testing used by other studies.^15,49^

Unlike our findings here, the Tanzanian study had not observed a decline in performance in older versus younger SCA participants.^27^ As that study tested children aged 6 years and older, taken together these findings support our findings of the sparing and/or resilience of the younger age group with SCA. Consistent with this observation, cumulative effects of SCA cerebrovascular injury over time are considered to be primarily responsible for the association between age and neurocognitive impairment in children with SCA.^50,51^ Collectively, these observations suggest that low caregiver education, alone and/or as a surrogate for low socioeconomic status (SES) and/or educational disadvantages may contribute to – but are not the main drivers of - the age effects seen on children with SCA in sub-Saharan Africa.^40^ Similar findings were reported in a U.S. study, where SCA and social factors both influenced neurocognition.^28^ The effect of age on neurocognitive outcome in our study could also be a consequence of “growing into deficit,” where effects of brain injury become apparent as the child grows older.^52^

A relatively high proportion of neurocognitive impairment in the older group of non-SCA siblings may be at least attributable to social issues, as all neurocognitively impaired siblings had caregivers with low education. Our group had previously reported this association among healthy children in Kampala.^40^

Study limitations include potential biases from SCA-associated survival and the cross-sectional study design. These issues may have affected the relationships seen with age. Nonetheless, our data reflect results from a substantial number of children receiving SCA care at a large urban center. Executive function was based on parental report rather than direct child assessment, hence could have been biased. More direct measures of executive function may provide clearer insights. A test for attention comparable to the TOVA for children ages 1-4 years was not available. Additional limitations include potential differential influences from illness-associated school absences adversely impacting test results and no assessment of attention in the younger age group.^53^ Contributions from hemoglobin concentration could not be discerned from SCA, as they are tightly linked. Unlike the random SCA clinic-based selection, sibling participation may have been biased, e.g. from possible parental concerns. Low caretaker education level in this sample may have had a downward effect on neurocognitive testing, although sibling assessment would have minimized those effects.^27,28^ No adjustments were made for multiple comparisons.

In conclusion, comparing a sample of Ugandan children with SCA to their non-SCA siblings aged 1-12 years, we demonstrated that: children with SCA had worse cognitive impairment and executive function than the unaffected siblings, and that these differences were attributable to the older age group, aged 5-12. The younger children with SCA were not different from their non-SCA siblings. Age 5-12 and prior stroke were most strongly associated with neurocognitive impairment, with some contribution from caregiver educational attainment. Low neurocognitive z-scores by age among non-SCA siblings suggests environmental influences, e.g., socio-economic status and education, among all participants, with potential parental selection bias for the siblings tested. Given the increased risk of impairment with age, interventions in early childhood may more likely provide benefit. Disease-modifying therapies, e.g. hydroxyurea, should be tested for stabilizing or improving neurocognitive function in young sub-Saharan children through amelioration of modifiable risk factors with SCA, including anemia.^54,55^

## Data Availability

All data produced in the present study are available upon reasonable request to the authors

## Acknowledgements

This work was supported by grants from the U.S. National Institutes of Health (NIH) (R21HD089791; Idro, Green) and R01HD096559, and acknowledges the NIH Fogarty program of Global Brain Disorders Research. We thank the outstanding Makerere-based BRAIN SAFE team and Global Health Uganda. Most of all, we thank the study participants and families for their contributions.

## Abbreviations

SCA: Sickle cell anemia
TCD: Transcranial doppler ultrasound
WHO: World Health Organization
ADHD: Attention-deficit/hyperactivity disorder
ANOVA: Analysis of Variance

## Acknowledgements

We acknowledge the parents and children who participated in the study and the outstanding study staff.

